# Vitamin D is a biomarker of clinical disease severity in oral lichen planus

**DOI:** 10.1101/2024.02.21.24303138

**Authors:** Sreedevi P. Unnikrishnan, Janice Boggon, Bernice Mclaughlin, Maggie E. Cruickshank, Rasha Abu-Eid, Karolin Hijazi

## Abstract

Oral lichen planus (OLP) is a chronic inflammatory condition known to adversely impact patient quality of life and is associated with an increased risk of cancer. The diverse clinical presentation and poor knowledge of clinical factors that determine the course of disease are amongst the main challenges that hinder effective and personalised treatment for OLP patients.

This study aimed to systematically identify clinical predictors of disease severity in OLP patients. A cohort of patients with histologically confirmed OLP (n=89) was recruited in a cross-sectional and single site study. A comprehensive assessment of clinical characteristics, medical and social history, haematological parameters, vitamin D levels, and Perceived Stress Scale (PSS-10) was carried out. Hierarchical linear regression identified the predictive value of clinical factors to OLP disease severity measured by the Oral Disease Severity Score (ODSS) and the Reticular/hyperkeratotic, Erosive/erythematous, Ulcerative (REU) scoring system.

Findings revealed that extraoral lichen planus and insufficient vitamin D levels were significant predictors of both overall and gingival disease severity of OLP. Specifically, patients with lichen planus affecting the skin or other mucosal sites had a 5.766-unit higher OLP severity score (β=5.766, 95% CI=.744-10.788, p=.025) than those without extraoral involvement as measured by ODSS. Patients with insufficient vitamin D levels exhibited 5.490-unit increase in OLP severity (β=5.490, 95% CI=1.136-9.844, p=.014) compared to those with adequate vitamin D levels. The presence of dental plaque-induced gingivitis (β=4.833, 95% CI=.974-8.692, p=.015), was found to be a significant factor affecting gingival disease severity.

This study revealed the importance of adequate vitamin D levels in OLP patients and suggests embedding vitamin D pre-treatment screening to optimise management of OLP. Future research should focus on elucidating the biological mechanisms underlying the protective effects of vitamin D in OLP.

## INTRODUCTION

Oral lichen planus (OLP) is a chronic inflammatory disease of the stratified squamous epithelium of the oral mucosa, characterised by distinctive clinical and histopathological features. The global prevalence of OLP is estimated at 1% (González-Moles et al. 2021). Women are more frequently affected than men, with a peak incidence in the fifth and sixth decades of life (Cheng et al. 2016). The pathogenesis and aetiology of OLP remain poorly understood, but it is thought to be a T-cell-mediated disorder (Cheng et al. 2016).

OLP presents with a spectrum of clinical subtypes with the most common clinical presentation being bilateral, symmetrical white reticular lesions on the posterior buccal mucosa (Olson, Rogers, Bruce 2016). Most cases of reticular OLP are asymptomatic, and incidentally identified by dental professionals (De Rossi and Ciarrocca 2014). In approximately 10% of cases, erosive or ulcerative lesions are confined to the gingiva, a clinical presentation known as desquamative gingivitis (Olson, Rogers, Bruce 2016). Erosive and atrophic OLP can cause varying degrees of pain (De Rossi and Ciarrocca 2014) and some studies have associated these forms with an increased risk of carcinoma (Olson, Rogers, Bruce 2016). The biological basis of inter-patient variability in the clinical presentation and course of disease of OLP is not well understood.

Therapeutic interventions for OLP evaluated in clinical trials, include topical and systemic corticosteroids, topical calcineurin inhibitors, retinoids and photochemotherapy, primarily with the goal of pain reduction (Lodi et al. 2012). However, these treatments are not effective in all OLP patients, notwithstanding that standardised measurement of efficacy is hampered by the lack of universal severity indicators (Unnikrishnan et al. 2023). Diversity of response to treatment is attributable to differences in host-related clinical and molecular factors.

While the exact cause of OLP remains elusive, several risk factors have been linked to the development of this chronic inflammatory condition. These include genetic predisposition (Gupta, S. and Jawanda 2015; Sun et al. 2000), potential involvement of infectious agents (Baek and Choi 2017), pre-existing connective tissue diseases (De Porras-carrique et al. 2022) and other systemic conditions like diabetes mellitus (Mallah et al. 2021; Otero Rey et al. 2018), thyroid diseases (Dave, Shariff, Philipone 2020; De Porras-carrique et al. 2022; Li et al. 2017), hyperlipidaemia (Lai, Yew, Schwartz 2016) and hypertension (De Porras-Carrique, Ramos-García, González-Moles 2023). Additionally, psychological factors like anxiety, stress (Chaudhary 2004; De Porras-Carrique et al. 2021) have been implicated in OLP progression. Despite these associations, there is little or no understanding of how these risk factors influence the course of disease. This study aimed to identify clinical predictors of disease severity in OLP to inform treatment optimisation and prognosis of OLP patients.

## MATERIALS AND METHODS

### Study population

Ethical approval for recruitment of OLP patients in this study was obtained from the West Midlands-Black Country Research Ethics Committee (Reference number: 21/WM/0247). Written informed consent was obtained from all participants in compliance with the Declaration of Helsinki. The study is registered at ClinicalTrials.gov. NCT05330572. This cross-sectional study was fully compliant with STROBE guidelines. Patients presenting with OLP diagnosed using Van der Meij and Van der Waal criteria (Van Der Meij and Van Der Waal 2003) at the Departments of Oral Medicine and Oral & Maxillofacial Surgery (Aberdeen Royal Infirmary) between December 2021 and December 2023 were eligible to the study. The following exclusion criteria were applied: age<18 years, current systemic steroid or immunosuppressive therapy for OLP, oral lichenoid contact reactions caused by dental materials, lichenoid drug reactions, graft-versus-host disease, other systemic inflammatory conditions associated with lichen planus-like oral lesions, isolated palatal lesions clinically consistent with Discoid Lupus Erythematosus, other concurrent oral mucosal inflammatory disorders or potentially malignant lesions, pregnancy or lactation.

### Study protocol

Participants’ medical and social history was recorded. Demographic and relevant clinical data (medical history, smoking status, and alcohol consumption) of study participants were collected from electronic patient records. If unavailable in electronic patient records, data were collected directly from patients during the study visit. The 10-item Perceived Stress Scale (PSS-10), a validated self-report instrument, was used to assess perceived stress levels in patients with OLP (Wiriyakijja et al. 2020). A single trained examiner carried out all oral examinations to assess OLP disease severity, oral hygiene status [Simplified Oral Hygiene Index (OHI-S)] (Greene and Vermillion 1964), number of teeth missing, and routine basic periodontal examination (Ower 2016). Gingivitis was diagnosed based on bleeding on probing sites (Chapple et al. 2018) and periodontitis based on amount of clinical attachment loss according to the 2017 classification system (Tonetti, Greenwell, Kornman 2018).Venous blood samples were collected for assessment of haematologic and biochemical parameters, including full blood count, haematinics, vitamin D. All parameters were assessed at the Laboratory Medicine diagnostic departments of Aberdeen Royal Infirmary and local reference ranges used.

### Assessment of clinical disease severity

The outcome measure was OLP disease severity measured as the Oral Disease Severity Score (ODSS) (Escudier et al. 2007) and Reticular/hyperkeratotic, Erosive/erythematous, Ulcerative (REU scoring system) (Piboonniyom et al. 2005).The ODSS is considered the most validated grading tool of disease activity in OLP, according to a recent systematic review (Unnikrishnan et al. 2023). ODSS assesses the site score and severity score of OLP-related lesions at seventeen oral mucosal subsites (Escudier et al. 2007). Gingival disease severity scores were calculated as the sum of site and activity scores (Escudier et al. 2007), confined to gingival sextants. The REU scoring system is one of the most reported scoring systems and has undergone the highest number of reliability assessments (Unnikrishnan et al. 2023).

### Statistical analyses

For descriptive statistics, normally distributed variables are presented as mean ± standard deviation (SD), and skewed variables as median and interquartile range [IQR]). Categorical variables are summarised as frequencies (n) and percentages (%).

We employed a two-phase analytical approach to identify significant predictors of OLP disease severity measured by ODSS. Initially, simple linear regression explored significant associations between clinical indicators and ODSS (p <0.05). Subsequently, hierarchical linear regression examined the independent contributions of selected clinically relevant indicators to ODSS, ODSS excluding pain score and REU scores. Covariate selection for the basic regression model employed a dual approach considering both evidence of existing literature and clinical relevance (Appendix Table 1). The basic regression model-I prioritised co-morbidities associated with OLP as established from systematic reviews, namely diabetes mellitus (Mallah et al. 2021; Otero Rey et al. 2018), hypothyroidism (De Porras-carrique et al. 2022; Li et al. 2017), hyperlipidaemia (Lai, Yew, Schwartz 2016) alongside clinically relevant demographic factors (age, gender) and important confounding factors (use of medications associated with lichenoid drug reactions, use of oral topical steroids within the past two months) as listed in Appendix Table 1. Subsequent models incorporated additional markers of interest individually: PSS-10 scores, extraoral manifestations of lichen planus, involvement of typically spared oral sites, vitamin D, vitamin B12, ferritin and serum folate. Each linear regression model quantified the incremental change (β) and 95% confidence intervals (CIs) in disease severity associated with a unit change in the respective marker. Statistical significance was set at p<0.05.

**Table 1:** General clinical characteristics of the study population.

|  | **Oral lichen planus population (n=89)** |
| --- | --- |
| Females, n (%) | 65 (73) |
| Age in years (median, [IQRs]) | 66 [58 to 73] |
| Oral Disease Severity Score (median, [IQRs]) | 10 [4 to 21] |
| Oral Disease Severity Score excluding pain score (median, [IQRs]) | 9 [4 to 19] |
| Gingival Oral Disease Severity Score (median, [IQRs]) | 1 [0 to 12] |
| Reticulation, erythematous and ulcerative score (median, [IQRs]) | 8 [4.5 to 20] |
| Pain score (median, [IQRs]) | .00 [.00 to 4.00] |
| Smoking pack years (median, [IQRs]) | .00 [.00 to 7.250] |
| Alcohol consumption (units per week) (median, [IQRs]) | 2.300 [.000 to 10.250] |
| Body Mass Index (kg/m<sup>2</sup>) (median, [IQRs]) | 26.290 [23.525 to 31.730] |
| 10 item-Perceived Stress Scale score (mean ± standard deviation) | 13.85±6.805 |
| Extraoral lichen planus, n (%) | 17 (19.1) |
| Genital lichen planus, n (%) | 12 (13.5) |
| Cutaneous lichen planus, n (%) | 5 (5.6) |
| Gingival involvement in oral lichen planus, n (%) | 46 (51.7) |
| Erosive clinical phenotype, n (%) | 24 (27) |
| Involvement of typically spared oral sites, n (%) | 19 (21.3) |
| Involvement of more than two oral subsites, n (%) | 56 (62.9) |
| Diabetes (type 2), under treatment n (%) | 12 (13.5) |
| Hypertension, under treatment n (%) | 25 (28.1) |
| Hyperlipidaemia, under treatment n (%) | 22 (24.7) |
| Coronary artery disease, n (%) | 9 (10.1) |
| Cerebrovascular disease, n (%) | 5 (5.6) |
| Inflammatory bowel disease, n (%) | 1 (1.1) |
| Connective tissue diseases (scleroderma, rheumatoid arthritis, Sjogren's syndrome), n (%) | 5 (5.6) |
| Scleroderma, n | 1 |
| Rheumatoid arthritis, n | 2 |
| Sjogren's syndrome, n | 2 |
| Other autoimmune diseases (psoriasis, vitiligo, alopecia areata, coeliac disease, pernicious anaemia, bullous pemphigoid, lichen sclerosus), n (%) | 11(12.4) |
| Psoriasis, n | 2 |
| Vitiligo, n | 1 |
| Alopecia areata, n | 1 |
| Coeliac disease, n | 4 |
| Pernicious anaemia, n | 3 |
| Bullous pemphigoid, n | 1 |
| Lichen sclerosus, n | 1 |
| Hypothyroidism, under treatment n (%) | 18 (20.2) |

| **Overall Oral Hygiene Index-Simplified, n (%)** |  |
| --- | --- |
| Edentulous/missing index teeth | 31 (34.8) |
| Good | 53 (59.6) |
| Fair | 5 (5.6) |
| Poor | 0 |
| Gingivitis ($\geq 10\%$ bleeding sites), n (%) | 17 (19.1) |
| Periodontitis, n (%) | 46 (51.7) |
| Recent use of oral topical steroids (within past two months), n (%) | 33 (37.1) |
| Coexisting fungal infection, n (%) | 5 (5.6) |
| Insufficient vitamin D levels (25-50 nmol/L), n (%) | 29 (32.6) |
| Deficient vitamin D levels ($< 25$ nmol/L), n (%) | 8 (9) |
| On replacement therapy for vitamin B12, n (%) | 18 (20.2) |
| Deplete vitamin B12 levels, n (%) | 13 (14.6) |
| On replacement therapy for ferritin, n (%) | 5 (5.6) |
| Deplete ferritin levels, n (%) | 4 (4.5) |
| On replacement therapy for folate, n (%) | 5 (5.6) |
| Deplete folate levels, n (%) | 3 (3.4) |

**Abbreviation:** IQR, Interquartile Range.

Similarly, for gingival lichen planus, we constructed a baseline linear regression model-I considering key confounders such as dental plaque-induced gingivitis and periodontitis alongside relevant co-variates (age, gender, systemic comorbidities and use of oral topical steroids). Subsequent models incorporated co-variates of interest individually (extraoral involvement, vitamin D, and haematinics) to explore their independent associations with gingival disease severity measured by the gingival ODSS. Each model again provided the β and 95% CI for the incremental change in gingival severity associated with a unit change in the respective covariate. Data were analysed using IBM SPSS Statistics v.29.0.1.0 (IBM, Hampshire, UK).

The sample size for hierarchical linear regression was determined a priori aiming for a statistical power of 0.90 to detect a medium effect size (f² = 0.15) for the effect of exposures of interest on the dependent variable. We considered exposures of interest as tested effects, while the remaining eight variables were included in the basic regression model. Based on this power calculation, a minimum sample size of 88 participants was required (G*Power version 3.1.9.7).

## RESULTS

### Characteristics of study participants

Eighty-Nine patients out of a total of 270 eligible OLP patients presenting consecutively to the Oral Medicine and Oral & Maxillofacial Surgery outpatient clinics between December 2021 and December 2023 agreed to participate and were enrolled into the study. Characteristics of study participants are reported in **Table *1***. The median [IQRs] age of the study population was 66 [58 to 73] years, and 65 (73%) patients were females. The cohort was representative of the typical OLP population: higher prevalence of females in post-menopausal age. Seventeen participants (88.2% females) presented with extraoral lichen planus, with a notable prevalence of genital involvement (over two-thirds). Nearly half (47.1%) of the patients in the cohort suffered from chronic diseases. Our cohort had a lower prevalence of vitamin D insufficiency (32.6%) and deficiency (9%) compared to global rates (47.9% and 15.7%, respectively) (Cui et al. 2023).

The median total ODSS score was 10 (range: 0-44), indicating a mild to moderate disease presentation. The median ODSS score, excluding the pain component, was 9 (range: 0-38), while the median Gingival ODSS score was 1 (range: 0-18). The median REU score was 8 (range: 0-49).

### Selection of clinically relevant co-variates

For exploratory analysis of clinical factors significantly associated with the severity of OLP, a simple linear regression analysis was conducted, with OLP severity assessed using the ODSS as the dependent variable (Appendix Table 1). Statistically significant clinical variables (p< 0.05) associated with OLP severity included diabetes mellitus, hyperlipidaemia, extraoral manifestations of lichen planus, erosive clinical phenotype, involvement of typically spared oral sites, recent use of oral topical steroids (within the past two months), dental plaque-induced gingivitis and insufficient vitamin D levels (Appendix Table 1). Additional co-variates were based on formal evidence of clinical characteristics associated with OLP or probable confounders according to accepted clinical knowledge and irrespective of statistical significance in exploratory analyses, namely age, gender, hypothyroidism, connective tissue diseases (scleroderma, rheumatoid arthritis, Sjogren’s syndrome), other autoimmune diseases (psoriasis, vitiligo, alopecia areata, coeliac disease, pernicious anaemia, bullous pemphigoid, lichen sclerosus), hypertension, coronary artery disease, inflammatory bowel disease, history of cancer, anxiety or depression, smoking, alcohol consumption, stress levels (PSS-10), medications associated with lichenoid drug reactions, amalgam restorations, co-existing candidal infection, vitamin D, vitamin B12, ferritin, and serum folate levels (Appendix Table 1).

### Independent predictors of disease severity in OLP

The basic model-I (age, gender, diabetes mellitus, hypothyroidism, hyperlipidaemia, use of medications associated with lichenoid drug reactions, erosive phenotype, use of oral topical steroids) explained 33.0% of the variance in OLP severity and was a statistically significant predictor of OLP severity as assessed by total ODSS (p=<.001) (**Table 2**). The analysis identified three significant associations between OLP severity and the following independent variables: hyperlipidaemia (β= 6.548, p=0.044); erosive phenotype (β=11.758, p=<.001) and oral topical steroid use (β=4.926, p=0.023) (**Table 2**).

**Table 2:**
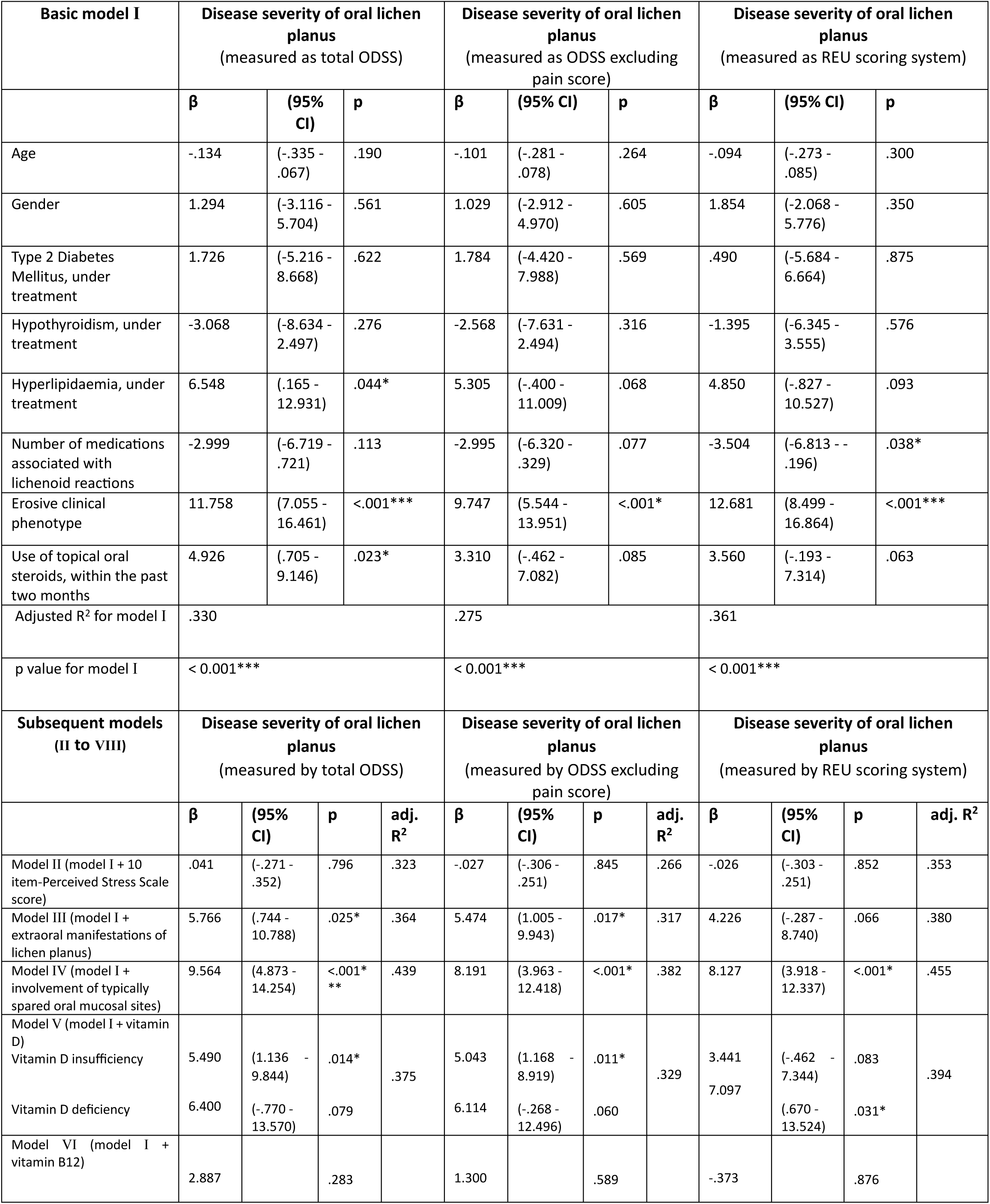
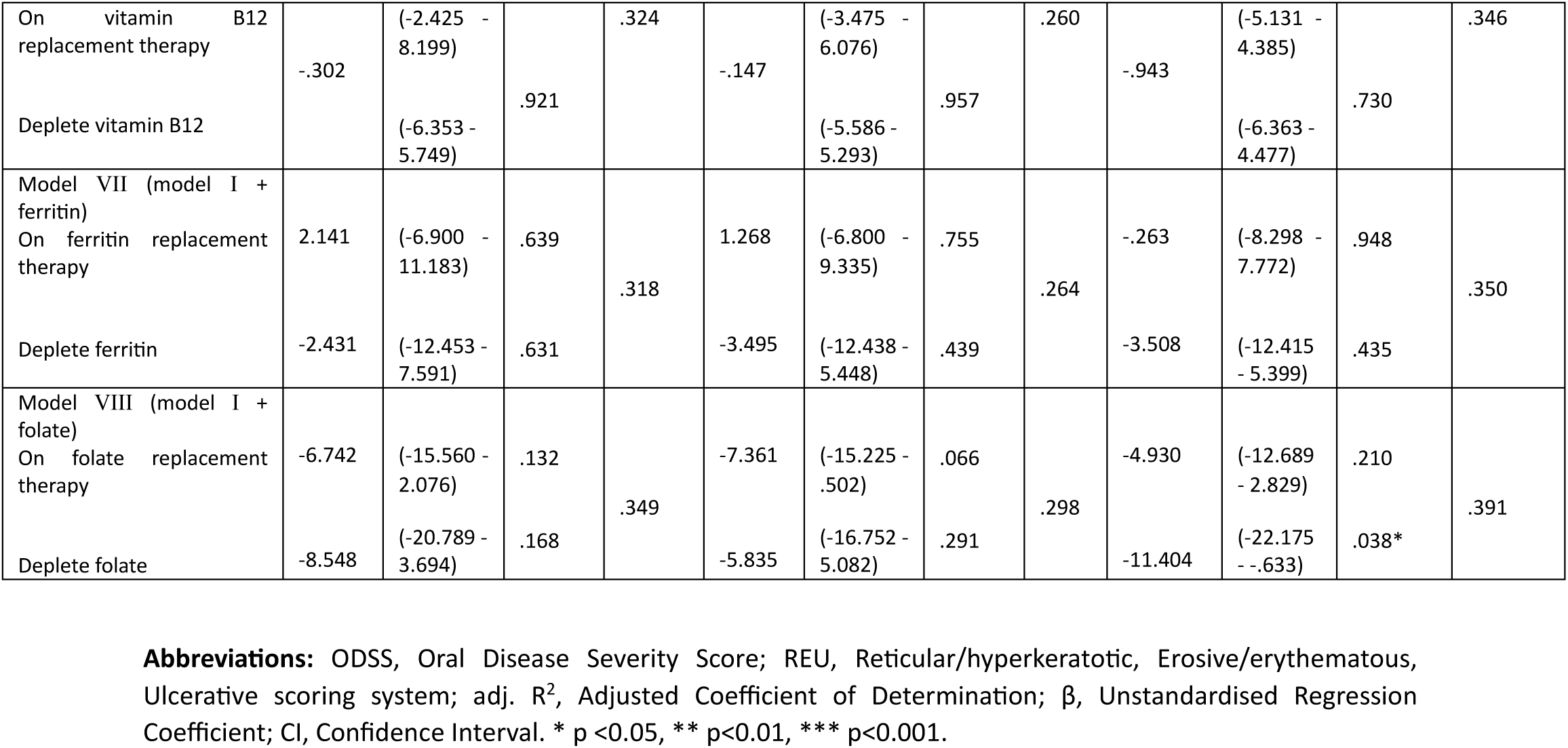
Hierarchical linear regression of clinical indicators of oral lichen planus disease severity.

Subsequently, after adjusting for basic model co-variates, hierarchical linear regression revealed that extraoral manifestations of lichen planus (model III, β=5.766, 95% CI=.744-10.788, p=.025), involvement of typically spared oral sites (model IV, β=9.564, 95% CI= 4.873-14.254, p=<.001) and insufficient vitamin D (model V, β=5.490, 95% CI=1.136-9.844, p=.014) were significant predictors of disease severity (**Table 2**). Specifically, patients with lichen planus affecting skin or other mucosal sites had a 5.766-unit higher OLP severity score than those without extraoral involvement. Similarly, individuals with OLP affecting less frequent oral subsites (palate, floor of the mouth and upper lip) (Olson, Rogers, Bruce 2016; Scully and Carrozzo 2007) had a 9.564-unit increase in OLP severity compared to those with more typical involvement (buccal mucosa, tongue). Finally, patients with insufficient vitamin D levels exhibited 5.490-unit increase in OLP severity compared to those with adequate vitamin D levels.

The predictive value of co-variates was assessed in relation to other measures of OLP severity (ODSS excluding pain score and REU scores) to test generalisability of findings to other grading tools. The basic model-I was a statistically significant predictor and explained 27.5% and 36.1% of the variance in OLP severity as assessed by ODSS excluding pain scores and REU scores, respectively. Subsequent hierarchical regression in relation to ODSS excluding pain scores as outcome and adjusted for confounders, revealed the same three independent predictors as regression in relation to total ODSS, namely extraoral manifestations of lichen planus (model III, β=5.474, 95% CI=1.005-9.943, p=.017), involvement of typically spared oral mucosal sites (model IV, β=8.191, 95% CI=3.963-12.418, p=<.001) and insufficient vitamin D levels (model V, β=5.043, 95% CI=1.168–8.919, p=.011) (**Table 2**). On the other hand, hierarchical regression in relation to REU scores, again adjusted for confounders, revealed less commonly involved oral sites (model IV, β=8.127, 95% CI=3.918-12.337, p=<.001), deficient vitamin D levels (model V, β= 7.097, 95% CI=.670-13.524, p=.031) and deplete folate levels (model VIII, β=-11.404, 95% CI=-22.175--.633, p=.038) (**Table 2**) as independent predictors.

### Independent predictors of gingival disease severity

Independent predictors of gingival disease severity were assessed separately in order to consider confounding factors specifically relevant to gingival lichen planus, namely dental plaque-induced gingivitis and periodontitis. A basic linear regression model-I (age, gender, diabetes mellitus, hypothyroidism, plaque-induced gingivitis, periodontitis, erosive phenotype, use of oral topical steroids) explained 10.0% of the variance of OLP severity as measured by gingival ODSS. In this model, dental plaque induced gingivitis (β=4.833, p=.015), and erosive phenotype (β=3.425, p=.049) were confirmed as significant predictors of gingival disease severity (**Table 3**). Following adjustment for basic model co-variates, hierarchical linear regression confirmed extraoral involvement (model II, β=5.052, 95% CI=1.072-9.032, p=.014), and insufficient vitamin D (model III, β=4.732, 95% CI=1.541-7.923, p=.004) as independent predictors (**Table 3**).

**Table 3:** Hierarchical linear regression of clinical indicators of gingival disease severity (n=89).

| Basic model I | Gingival disease severity of oral lichen planus<br>(measured as Gingival ODSS) |  |  |  |
| --- | --- | --- | --- | --- |
|  | β | (95% CI) | p |  |
| Age | -.024 | (-.170 - .122) | .746 |  |
| Gender | .242 | (-3.086 - 3.569) | .885 |  |
| Type 2 Diabetes Mellitus, under treatment | 2.536 | (-2.172 – 7.243) | .287 |  |
| Hypothyroidism, under treatment | -1.919 | (-5.845 - 2.006) | .334 |  |
| Dental plaque induced gingivitis based on bleeding on probing | 4.833 | (.600 - 8.333) | .024* |  |
| Presence of periodontitis | 1.388 | (-1.676 - 4.452) | .370 |  |
| Erosive clinical phenotype | 3.425 | (.021 – 6.830) | .049* |  |
| Use of topical oral steroids, within the past two months | 1.141 | (-1.965 - 4.247) | .467 |  |
| Adjusted R<sup>2</sup> for model I | .100 |  |  |  |
| p value for model I | .034* |  |  |  |
| Subsequent models<br>(II to VI) | Gingival disease severity of oral lichen planus<br>(measured as Gingival ODSS) |  |  |  |
|  | β | (95% CI) | p | adj. R<sup>2</sup> |
| Model II (model I + extraoral manifestations of lichen planus) | 5.052 | (1.072 - 9.032) | .014* | .157 |
| Model III (model I + vitamin D) |  |  |  |  |
| Vitamin D insufficiency | 4.732 | (1.541 - 7.923) | .004** | .176 |
| Vitamin D deficiency | 3.842 | (-1.385 - 9.070) | .147 |  |
| Model IV (model I + vitamin B12) |  |  |  |  |
| On vitamin B12 replacement therapy | 1.349 | (-2.633 - 5.332) | .502 | .083 |
| Deplete vitamin B12 | -.301 | (-4.727 - 4.125) | .893 |  |
| Model V (model I + ferritin) |  |  |  |  |
| On ferritin replacement therapy | 2.488 | (-4.257 - 9.232) | .465 | .089 |
| Deplete ferritin | -2.582 | (-10.179 - 5.014) | .501 |  |
| Model VI (model I + folate) |  |  |  |  |
| On folate replacement therapy | -4.677 | (-11.391 - 2.038) | .169 | .103 |
| Deplete folate | -3.072 | (-12.436 - 6.292) | .516 |  |

**Abbreviations:** ODSS, Oral Disease Severity Score; adj. R<sup>2</sup>, Adjusted Coefficient of Determination; β, Unstandardised Regression Coefficient; CI, Confidence Interval. \* p <0.05, \*\* p<0.01, \*\*\* p<0.001.

## DISCUSSION

The diverse clinical presentation and poor knowledge of clinical factors that determine the course of OLP are amongst the main challenges that hinder effective and personalised treatment. In this cross-sectional study of OLP patients presenting with a spectrum of clinical severity, we revealed that extraoral manifestations of lichen planus and vitamin D were consistent clinical predictors of both overall and gingival disease severity of OLP.

The significant association between extraoral lichen planus and increased OLP severity identified in our study supports the notion that multi-site disease contributes to greater oral involvement. In our study, genital involvement, particularly vaginal-vulval lichen planus, was more prevalent than cutaneous involvement. This finding warrants further investigation to formally determine if predilection for certain extraoral sites influences OLP severity. Further, the involvement of typically spared oral mucosal sites (floor of the mouth, palate and upper lip) (Olson, Rogers, Bruce 2016; Scully and Carrozzo 2007) was a significant predictor of OLP severity. This finding suggests that the distribution of lesions to atypical mucosal sites may be associated with a more severe disease course. The involvement of atypical oral sites and, more broadly, multi-site involvement could reflect more profound T-cell-mediated immune dysregulation in OLP resulting in more severe disease manifestations.

The relationship between vitamin D status and OLP severity supports the findings of previous reports of lower vitamin D levels in OLP patients compared to controls in a range of geographical settings (Aksu Arıca et al. 2020; Bahramian et al. 2018; Gupta, A. et al. 2017; Muzaffar Tak and Hussain Chalkoo 2017). Further, lower vitamin D levels were reported in erosive OLP patients compared to non-erosive types, albeit not consistently at statistical significance (Aksu Arıca et al. 2020; Gupta, A. et al. 2017). In this study, we have demonstrated the independent role of vitamin D as a predictor of clinical severity accounting for a range of potential confounding variables associated with OLP.

Vitamin D is a pleiotropic hormone that exerts a wide range of physiological functions by binding to the vitamin D receptor (VDR) (Saeed et al. 2022). In vitro studies have shown that vitamin D/VDR signalling protects against OLP by suppressing inflammation and epithelial cell apoptosis (Du et al. 2017; Zhao et al. 2018; Zhao et al. 2019). In this study, we showed for that vitamin D was a robust predictor of OLP disease severity, suggesting that insufficient vitamin D levels may contribute to disease progression. Vitamin D exhibits anti-inflammatory properties through downregulation of Th1 and upregulation of Th2 responses (Saeed et al. 2022). Vitamin D/VDR signalling also suppresses apoptosis in oral keratinocytes by repressing miR-802 expression (Zhao et al. 2019). Vitamin D deficiency in OLP may compromise the protective effect of vitamin D/VDR signalling, resulting in increased apoptosis of oral keratinocytes and contributing to the development of erosions or ulcerations seen in more severe clinical presentations.

In patients with gingival manifestations of OLP, we showed a significant association between gingival disease severity and plaque-induced gingivitis. This observation aligns with accepted knowledge that pain associated with gingival OLP interferes with effective oral hygiene, leading to plaque accumulation and in turn exacerbating existing OLP lesions and vice versa (Scribante et al. 2023). Extraoral lichen planus and insufficient vitamin D emerged as significant predictors of gingival disease severity in our study. This finding suggests a potential interplay between systemic and local factors influencing gingival health in OLP patients. It highlights the need for a holistic approach addressing both local and systemic factors, for example vitamin D supplementation, in optimising oral health outcomes of OLP patients.

Statistically significant relationships between haematinic factors and disease severity were not consistently identified across all outcomes of disease severity used in this study. Specifically, we observed a significant association between folate deficiency and OLP severity only when this was measured by the REU score. This finding questions the role of haematinic deficiencies as a predictor of disease severity, notwithstanding the limitations of this single arm cross-sectional study. Further, our analysis did not identify a significant association between stress and OLP severity. This could be attributed to the inherent limitations of self-reported stress data, which are susceptible to recall bias. Likewise, we observed a statistically significant association between hyperlipidaemia and OLP severity as measured by ODSS, but the associated wide confidence intervals suggest data uncertainty.

This study highlights the importance of adequate vitamin D supplementation in OLP in the population here described. Vitamin D deficiency is a global public health concern, affecting individuals of all ages, including low-latitude regions with ample sunlight (Palacios and Gonzalez 2015). On the other hand, geographical differences in the prevalence of vitamin D deficiency partly dependent on sunlight exposure are well documented, for example in the United Kingdom where prevalence is higher in the northern regions (Lin et al. 2021). To date, no studies have formally assessed the relationship between sunlight exposure, the subsequent impact on vitamin D status, and the geographical distribution of oral lichen planus. A recent systematic review (González-Moles et al. 2021) reported the highest prevalence of OLP in Europe compared to the global population. However, in nation-level analysis higher OLP prevalences in European countries did not align with reduced sunlight exposure.

Future research should explore generalisability of the findings of this study to the wider population as well as the biological mechanisms underlying the beneficial effects of vitamin D in OLP. Furthermore, interventional studies are required to assess the therapeutic efficacy of vitamin D supplementation in OLP to determine its clinical benefit and to optimise treatment guidelines.

## Acknowledgement

We extend our sincere gratitude to the clinical and administrative staff of the Departments of Oral Medicine and Oral & Maxillofacial Surgery at Aberdeen Royal Infirmary for their invaluable assistance with patient recruitment and research visits. We are further indebted to all the patients who generously participated, making this research possible. We also thank the Medical Statistics team at the University of Aberdeen for their guidance with the statistical analysis presented in the study.

## Author contributions

Sreedevi P. Unnikrishnan: contributed to data acquisition, analysis or interpretation of data and drafted the manuscript; Janice Boggon: contributed to design and critically revised the manuscript; Bernice Mclaughlin: contributed to design and critically revised the manuscript; Maggie E. Cruickshank: contributed to conception, design, analysis or interpretation of data and critically revised the manuscript; Rasha Abu-Eid: contributed to conception, design, analysis or interpretation of data and critically revised the manuscript; Karolin Hijazi: contributed to conception, design, analysis or interpretation of data and critically revised the manuscript. All authors gave their final approval and agree to be accountable for all aspects of the work.

## Data availability statement

Pseudonymised data can be made available upon reasonable and FAIR-compliant requests to the corresponding author.

## References

1. Aksu Arıca D, Baykal Selçuk L, Örem A, Karaca Ural Z, Yaylı S, Bahadır S. 2020. Evaluation of serum vitamin D levels in patients with lichen planus. Turkderm. 54(4):138–142.

2. Baek K, Choi Y. 2017. The microbiology of oral lichen planus: Is microbial infection the cause of oral lichen planus? Mol Oral Microbiol. 33(1):22–28.

3. Bahramian A, Bahramian M, Mehdipour M, Falsafi P, Khodadadi S, Dabaghi Tabriz F, Deljavanghodrati M. 2018. Comparing vitamin D serum levels in patients with oral lichen planus and healthy subjects. J Dent (Shiraz).19(3):212–216.

4. Chapple ILC, Mealey BL, Dyke TE, Bartold PM, Dommisch H, Eickholz P, Geisinger ML, Genco RJ, Glogauer M, Goldstein M, et al. 2018. Periodontal health and gingival diseases and conditions on an intact and a reduced periodontium: Consensus report of workgroup 1 of the 2017 world workshop on the classification of periodontal and Peri-Implant diseases and conditions. J. Periodontol. 89(S1):S74–84.

5. Chaudhary S. 2004. Psychosocial stressors in oral lichen planus. Aust Dent J. 49(4):192–195.

6. Cheng YL, Gould A, Kurago Z, Fantasia J, Muller S. 2016. Diagnosis of oral lichen planus: A position paper of the american academy of oral and maxillofacial pathology. Oral Surg Oral Med Oral Radiol.122(3):332–54.

7. Cui A, Zhang T, Xiao P, Fan Z, Wang H, Zhuang Y. 2023. Global and regional prevalence of vitamin D deficiency in population-based studies from 2000 to 2022: A pooled analysis of 7.9 million participants. Front Nutr (Lausanne). 10:1070808.

8. Dave A, Shariff J, Philipone E. 2020. Association between oral lichen planus and systemic conditions and medications: Case–control study. Oral Dis. 27(3):515–524.

9. De Porras-Carrique T, Ramos-García P, González-Moles MÁ. 2023. Hypertension in oral lichen planus: A systematic review and meta-analysis. Oral Dis [epub ahead of print].

10. De Porras-carrique T, Ramos-garcía P, Aguilar-diosdado M, Warnakulasuriya S, González-moles MÁ. 2022. Autoimmune disorders in oral lichen planus: A systematic review and meta-analysis. Oral Dis. 29(4):1382–1394.

11. De Porras-Carrique T, González-Moles MÁ, Warnakulasuriya S, Ramos-García P. 2021. Depression, anxiety, and stress in oral lichen planus: A systematic review and meta-analysis. Clin Oral Investig. 26(2):1391–1408.

12. De Rossi SS, Ciarrocca K. 2014. Oral lichen planus and lichenoid mucositis. Dent Clin N Am.58(2):299–313.

13. Du J, Li R, Yu F, Yang F, Wang J, Chen Q, Wang X, Zhao B, Zhang F. 2017. Experimental study on 1,25(OH) 2 D 3 amelioration of oral lichen planus through regulating NF- κ B signaling pathway. Oral Dis. 23(6):770–778.

14. Escudier M, Ahmed N, Shirlaw P, Setterfield J, Tappuni A, Black MM, Challacombe SJ. 2007. A scoring system for mucosal disease severity with special reference to oral lichen planus. Br J Dermatol.157(4):765–70.

15. González-Moles MÁ, Warnakulasuriya S, González-Ruiz I, González-Ruiz L, Ayén Á, Lenouvel D, Ruiz-Ávila I, Ramos-García P. 2021. Worldwide prevalence of oral lichen planus: A systematic review and meta-analysis. Oral Dis. 27(4):813–828.

16. Greene JC, Vermillion JR. 1964. The simplified oral hygiene index. J Am Dent Assoc. 68(1):7–13.

17. Gupta A, Sasankoti Mohan RP, Kamarthi N, Malik S, Goel S, Gupta S. 2017. Serum vitamin D level in oral lichen planus patients of north India-A case-control study. J Dermatol Res Ther. 1(2):19–35.

18. Gupta S, Jawanda MK. 2015. Oral lichen planus: An update on etiology, pathogenesis, clinical presentation, diagnosis and management. Indian J Dermatol. 60(3):222–9.

19. Lai YC, Yew YW, Schwartz RA. 2016. Lichen planus and dyslipidemia: A systematic review and meta-analysis of observational studies. Int J Dermatology. 55(5): e295–304.

20. Li D, Li J, Li C, Chen Q, Hua H. 2017. The association of thyroid disease and oral lichen planus: A literature review and meta-analysis. Front Endocrinol (Lausanne). 8:310.

21. Lin L, Smeeth L, Langan S, Warren-Gash C. 2021. Distribution of vitamin D status in the UK: A cross-sectional analysis of UK biobank. BMJ Open. 11(1):e038503.

22. Lodi G, Carrozzo M, Furness S, Thongprasom K. 2012. Interventions for treating oral lichen planus: A systematic review. Br J Dermatol. 166(5):938–47.

23. Mallah N, Ignacio Varela-centelles P, Seoane-romero J, Takkouche B. 2021. Diabetes mellitus and oral lichen planus: A systematic review and meta-analysis. Oral Dis. 28(8):2100–2109.

24. Muzaffar Tak M, Hussain Chalkoo A. 2017. Vitamin d deficiency-a possible contributing factor in the aetiopathogenesis of oral lichen planus. J Evolution Med Dent Sci. 6(66):4769–4772.

25. Olson MA, Rogers RS, Bruce AJ. 2016. Oral lichen planus. Clin Dermatol. 34(4):495–504.

26. Otero Rey EM, Yáñez-busto A, Rosa Henriques IF, López-lópez J, Blanco-carrión A. 2018. Lichen planus and diabetes mellitus: Systematic review and meta-analysis. Oral Dis. 25(5):1253–1264.

27. Ower P. 2016. BPE guidelines: British society of periodontology revision 2016. Dent Update. 43(5):406–8.

28. Palacios C, Gonzalez L. 2015. Is vitamin D deficiency a major global public health problem? J Steroid Biochem Mol Biol.144:138–45.

29. Piboonniyom S, Treister N, Pitiphat W, Woo S. 2005. Scoring system for monitoring oral lichenoid lesions: A preliminary study. Oral Surg Oral Med Oral Pathol Oral Radiol Endod. 99(6):696–703.

30. Saeed S, Choudhury P, Ahmad SA, Alam T, Panigrahi R, Aziz S, Kaleem SM, Priyadarshini SR, Sahoo PK, Hasan S. 2022. Vitamin D in the treatment of oral lichen planus: A systematic review. Biomedicines. 10(11):2964.

31. Scribante A, Pellegrini M, Li Vigni G, Pulicari F, Spadari F. 2023. Desquamative gingivitis, oral hygiene, and autoimmune oral diseases: A scoping review. Appl Sci. 13(18):10535.

32. Scully C, Carrozzo M. 2007. Oral mucosal disease: Lichen planus. Br J Oral Maxillofac Surg. 46(1):15–21.

33. Sun A, Wu YC, Wang JT, Liu BY, Chiang CP. 2000. Association of HLA-te22 antigen with anti-nuclear antibodies in chinese patients with erosive oral lichen planus. Proc Natl Sci Counc Repub China B. 24(2):63–9.

34. Tonetti MS, Greenwell H, Kornman KS. 2018. Staging and grading of periodontitis: Framework and proposal of a new classification and case definition. J Periodontol. 89(S1):S159–72.

35. Unnikrishnan SP, Rampersaud E, Mcgee A, Cruickshank ME, Abu-Eid R, Hijazi K. 2023. Disease severity scoring systems in mucosal lichen planus: A systematic review. Oral Dis. 29(8):3136–51.

36. Van Der Meij EH, Van Der Waal I. 2003. Lack of clinicopathologic correlation in the diagnosis of oral lichen planus based on the presently available diagnostic criteria and suggestions for modifications. J Oral Pathol Med. 32(9):507–12.

37. Wiriyakijja P, Porter S, Fedele S, Hodgson T, McMillan R, Shephard M, Ni Riordain R. 2020. Validation of the HADS and PSS-10 and psychological status in patients with oral lichen planus. Oral Dis. 26(1):96–110.

38. Zhao B, Li R, Yang F, Yu F, Xu N, Zhang F, Ge X, Du J. 2018. LPS-induced vitamin D receptor decrease in oral keratinocytes is associated with oral lichen planus. Sci Rep. 8(1):763–9.

39. Zhao B, Xu N, Li R, Yu F, Zhang F, Yang F, Ge X, Li YC, Du J. 2019. Vitamin D/VDR signaling suppresses microRNA-802-induced apoptosis of keratinocytes in oral lichen planus. FASEB J. 33(1):1042–1050.

