## Appendix Table 1 for "Vitamin D is a biomarker of clinical disease severity in oral lichen planus"

Appendix Table 1: Simple linear regression of clinical indicators of disease severity with Oral Disease Severity Score as outcome variable (n=89) along with the relevance of each variable judged by formal literature or accepted clinical knowledge.

| **Independent variables** | **Disease severity of oral lichen planus** (n=89)  **(measured as ODSS)** | | |  | **Relevance of variables supported by literature and clinical knowledge** | **Citations supporting variable relevance** |
| --- | --- | --- | --- | --- | --- | --- |
|  | **β** | **(95% CI)** | **p** | **R^2^** |  |  |
| Age | -.043 | (-.260 - .174) | .694 | .002 | +++ | (González‐Moles et al. 2021) |
| Gender  (female vs male) | 1.540 | (-3.835 - 6.914) | .571 | .004 | +++ | (González‐Moles et al. 2021) |
| Type 2 Diabetes Mellitus, under treatment  (yes vs no) | 7.080 | (.248 - 13.912) | .042* | .046 | +++ | (Mallah et al. 2021; Otero Rey et al. 2018) |
| Hyperlipidaemia, under treatment  (yes vs no) | 7.936 | (2.660 - 13.211) | .004* | .093 | +++ | (Lai, Yew, Schwartz 2016) |
| Hypothyroidism, under treatment  (yes vs no) | -.191 | (-6.140 - 5.758) | .949 | .000 | +++ | (De Porras‐carrique et al. 2022; Li et al. 2017) |
| Connective tissue diseases (scleroderma, rheumatoid arthritis, Sjogren’s syndrome)  (yes vs no) | .733 | (-9.643 - 11.110) | .889 | .000 | ++ | (De Porras‐carrique et al. 2022) |
| Other autoimmune diseases: psoriasis, vitiligo, alopecia areata, coeliac disease, pernicious anaemia, bullous pemphigoid, lichen sclerosus  (yes vs no) | -1.638 | (-8.890 - 5.615) | .655 | .002 | ++ | (De Porras‐carrique et al. 2022) |
| Hypertension, under treatment  (yes vs no) | 1.129 | (-4.182 - 6.441) | .674 | .002 | +++ | (De porras‐carrique, Ramos‐garcía, González‐moles 2023) |
| h/o coronary artery disease  (yes vs no) | 5.269 | (-2.577 - 13.116) | .185 | .020 | ++ | (Conrotto et al. 2018) |
| Inflammatory bowel disease  (yes vs no) | 7.375 | (-15.242 - 29.992) | .519 | .005 | ++ | (De Porras‐carrique et al. 2022) |
| h/o of cancer (any tissue origin)  (yes vs no) | 1.397 | (-5.593 - 8.388) | .692 | .002 | ++ | (Dave, Shariff, Philipone 2020) |
| Anxiety/depression, under treatment  (yes vs no) | .773 | (-5.057 - 6.602) | .793 | .001 | +++ | (De Porras-Carrique et al. 2021) |
| Smoking in pack years | -.047 | (-.216 - .122) | .580 | .004 | ++ | (Kłosek et al. 2011) |
| Alcohol consumption in units per week | -.010 | (-.233 - .213) | .931 | .000 | + | (Xu et al. 2009) |
| 10 item-Perceived Stress Scale score | .118 | (-.234 - .470) | .508 | .005 | +++ | (De Porras-Carrique et al. 2021) |
| Extraoral manifestations of lichen planus  (presence vs absence) | 9.087 | (3.324 - 14.849) | .002* | .101 | Clinical observation | - |
| Erosive clinical phenotype  (erosive vs non-erosive) | 13.294 | (8.714 - 17.873) | <.001* | .277 | Clinical observation | - |
| Involvement of typically spared oral sites  (presence vs absence) | 13.688 | (8.638 - 18.738) | <.001* | .250 | Clinical observation | - |
| Number of amalgam restorations | .028 | (-.613 - .669) | .931 | .000 | Probable confounder | - |
| Number of medications associated with lichenoid reactions | 1.598 | (-2.476 - 5.673) | .438 | .007 | Potential confounder | - |
| Use of oral topical steroids (within the past two months)  (yes vs no) | 6.773 | (2.041 - 11.505) | **.**006* | .085 | Probable confounder | - |
| Co-existing candidal infection  (yes vs no) | 8.786 | (-1.422 - 18.993) | **.**091 | **.**033 | Probable confounder | - |
| Dental plaque-induced gingivitis  (≥10% bleeding site vs <10% bleeding site) | 6.978 | (1.084 - 12.872) | .021* | **.**060 | + | (Scribante et al. 2023) |
| Vitamin D (deficiency vs insufficient vs adequate)  Vitamin D insufficiency  Vitamin D deficiency | 8.328  5.500 | (3.396 - 13.260)  (-2.582 - 13.582) | .001*  .180 | .119 | ++ | (Jaafari-Ashkavandi et al.2023; Muzaffar Tak and Hussain Chalkoo 2017) |
| Vitamin B12 (on replacement vs no deficiency vs deplete)  On replacement therapy for vitamin B12  Deplete vitamin B12 | 2.663  -1.910 | (-3.410 - 8.735)  (-8.816 – 4.997) | .386  .584 | .015 | ++ | (Bao et al. 2020; Chen et al. 2015) |
| Ferritin (on replacement vs no deficiency vs deplete)  On replacement therapy for ferritin  Deplete ferritin | 1.525  3.275 | (-8.906 - 11.956)  (-8.318 - 14.868) | .772  .576 | .004 | ++ | (Bao et al. 2020; Chen et al. 2015) |
| Folate (on replacement vs no deficiency vs deplete)  On replacement therapy for folate  Deplete folate | -7.163  4.370 | (-17.469 - 3.143)  (-8.779 - 17.519) | .171  .511 | .028 | ++ | (Bao et al. 2020; Chen et al. 2015) |

**Abbreviations**: R^2^, Coefficient of Determination; β, Unstandardised Regression Coefficient; CI, Confidence Interval. *= statistically significant (p<0.05); +++ = covariate supported by systematic reviews; ++ = covariate supported by observational studies; + = covariate supported by opinions and qualitative data.
